# Focal surgical lesions reveal a subthalamic substrate for human wakefulness

**DOI:** 10.64898/2026.11.21.26368668

**Authors:** Shiro Horisawa, Atsushi Fukui, Kilsoo Kim, Takaomi Taira, Satoru Miyao

**Affiliations:** Department of Neurosurgery, Tokyo Women’s Medical University, Tokyo, Japan; Department of Neurosurgery, FJK Tokyo Hospital, Tokyo, Japan; Department of Neurosurgery, TMG Asaka Medical Center, Saitama, Japan

## Abstract

How focal subthalamic injury disrupts a distributed arousal system remains incompletely understood. We investigated transient somnolence after unilateral pallidothalamic tractotomy without sedation in 56 patients, including 14 with somnolence. Volume-adjusted probabilistic lesion mapping identified a hotspot dorsal to the lateral hypothalamus (MNI −6.0, −10.5, −4.0 mm; family-wise error-corrected P = 0.011). A model combining peak involvement and log-transformed lesion volume achieved a leave-one-out cross-validated area under the receiver operating characteristic curve (AUC) of 0.799. Applied without refitting to two external cohorts comprising 19 patients, it achieved an AUC of 0.830 (95% confidence interval 0.636–0.977; two-sided cohort-stratified exact permutation P = 0.0166). In a supplementary analysis, somnolence-associated lesions had greater map-weighted burden within a published structural connectivity map of consciousness recovery after deep brain stimulation (P = 0.0299). These findings identify a focal subthalamic vulnerability in maintaining human wakefulness, with a lesion-based model discriminating transient somnolence in independent surgical patients.

## Introduction

Wakefulness is supported by ascending pathways linking brainstem arousal nuclei with the hypothalamus, thalamus and basal forebrain.[1] This distributed organization raises an anatomical question: where can a focal injury impair the maintenance of wakefulness in humans? In thalamic stroke, severe arousal impairment is associated with lesion extension into the brainstem, illustrating the importance of distinguishing neighbouring structures.[2] Focal surgical interventions offer an opportunity to examine this question by comparing closely situated lesions with different clinical outcomes.

Pallidothalamic tractotomy (PTT) at Forel’s field interrupts pallidal efferent fibres to relieve refractory motor symptoms.[3] The procedure can also produce an abrupt, transient reduction in wakefulness. Gallay and colleagues described six cases of postoperative drowsiness or confusion that resolved within a day.[4] Godinho and colleagues reported somnolence in four of 12 patients after unilateral campotomy, resolving over 1–4 days.[5] In our previous series of cervical dystonia, two of 35 patients developed somnolence immediately after lesioning and recovered by the following morning.[6] PTT has also been associated with changes in sleep architecture and EEG activity.[7] These observations make the subthalamic surgical territory relevant to the study of human arousal.

The hypothalamus and its immediate surroundings are of particular interest. In humans, orexin/hypocretin neurons occupy the perifornical region of the posterior hypothalamus and extend into the lateral hypothalamus, with projections to multiple arousal-related structures.[8] Their marked depletion in human narcolepsy supports the importance of hypothalamic mechanisms in maintaining wakefulness.[9] Focal unilateral hypothalamic injury can also produce transient symptoms. Matsubara and colleagues described sudden sleep attacks and daytime sleepiness following a small unilateral hypothalamic infarction, with resolution the next day.[10] However, the spatial distribution of PTT lesions associated with somnolence remains poorly defined. Patients undergoing the same focal procedure permit comparison within this compact anatomical neighbourhood, while awake surgery without sedation allows observation of changes around the time of lesioning.

We used volume-adjusted probabilistic lesion mapping to identify the subthalamic location associated with transient somnolence after unilateral PTT. We then evaluated whether a model combining involvement of the statistical peak with lesion volume discriminated outcomes in internal cross-validation and separate validation cohorts. To place the lesion findings in an anatomical context, a supplementary analysis quantified weighted overlap with an independently published structural connectivity map associated with recovery of consciousness after deep brain stimulation.[11]

## Methods

### Study design and patients

We conducted a retrospective study of patients with movement disorders who underwent unilateral pallidothalamic tractotomy (PTT) at Tokyo Women’s Medical University between 2017 and 2022. Patients were included if the presence or absence of somnolence could be determined from the perioperative medical records and a postoperative lesion mask was available. Patients with insufficient documentation to determine somnolence status were excluded. The development cohort comprised 56 patients, each contributing one lesion mask. Validation was performed in two separate cohorts: 13 patients with movement disorders who underwent PTT at FJK Tokyo Hospital and six patients with epilepsy who underwent PTT at Tokyo Women’s Medical University. These cohorts enabled evaluation across an external centre and a different surgical indication at the development centre. Development procedures were performed by S.H.; the epilepsy and FJK validation procedures were performed by Dr Miyao and Dr Taira, respectively.

Ethics approval and consent: [committee names, approval numbers, and consent or opt-out arrangements].

### Clinical assessment of somnolence

Across the development and validation cohorts, procedures were performed with patients awake and without systemic sedation. Somnolence was assessed intraoperatively, immediately after surgery and during morning and evening rounds. Patients were classified as having somnolence when a new reduction in wakefulness was evident after lesioning, manifested by marked subjective sleepiness or reduced responsiveness to verbal stimulation. Assessment combined patient reports and bedside observation, using the same basic clinical approach across cohorts. Validation outcomes were ascertained by the respective treating teams. The presence or absence of somnolence was analysed as a binary outcome. Representative video recordings documented interaction before lesioning, the subsequent reduction in arousal and recovery.

### Lesion imaging and anatomical localization

Across all cohorts, lesions were manually delineated using MRIcroGL as the hyperintense region on T1-weighted MRI obtained within 24 h after surgery. Spatial normalization to MNI152NLin2009bAsym space used SynthMorph within Lead-DBS.[35] Binary lesion masks were analysed on a 0.5-mm isotropic grid, with right-sided lesions reflected across the midsagittal plane to align all lesions to the left hemisphere. Lesion volume was calculated as the number of non-zero mask voxels multiplied by the voxel volume.

Statistical and probability maps were overlaid on the hypothalamic-region atlas of Neudorfer and colleagues.[12] The spatial relationship between the volume-adjusted statistical peak and the lateral hypothalamus was assessed using the Euclidean distance in MNI space from the peak to the nearest atlas voxel centre.

### Volume-adjusted probabilistic lesion mapping

Voxel-wise analyses were restricted to locations included in the lesions of at least five development patients. At each eligible voxel, somnolence was modelled using lesion involvement at that voxel and total lesion volume. Volume was transformed using the natural logarithm and standardized to its mean and standard deviation. Firth bias-reduced logistic regression was used to address sparse lesion–outcome combinations,[36] followed by an intercept-only correction using Firth logistic regression with intercept correction (FLIC).[37] Continuous probability maps represented the fitted probability of somnolence with the voxel lesioned at the median lesion volume of the development cohort.

Inference on the spatial association was based on an efficient score test for voxel involvement under a logistic null model containing log-transformed lesion volume. A positive score indicated a greater likelihood of somnolence with involvement of that voxel. Family-wise error (FWER) across the search volume was controlled using 20,000 parametric bootstrap replicates. In each replicate, binary outcomes were simulated from the fitted volume-only null model while retaining the lesion masks and volumes. The null model was refitted, score statistics were recalculated, and the maximum positive score was retained. Corrected voxel-wise P values were calculated from this maximum-statistic distribution with a plus-one correction. The voxel with the largest observed positive volume-adjusted score defined the statistical peak.

Supporting analyses comprised unadjusted voxel-wise Fisher exact tests with FWER correction using 20,000 label permutations, a two-sided analysis using the maximum absolute score, and repetition of the adjusted mapping after exclusion of two cases with atypical lesion locations identified during spatial quality review.

### Prediction model and internal validation

The peak-plus-volume model combined a binary indicator of whether a lesion included the volume-adjusted statistical peak with standardized log-transformed lesion volume. Model coefficients were estimated using Firth logistic regression followed by FLIC. A volume-only model was evaluated as a reference using the same fitting approach.

Internal discrimination was evaluated using leave-one-out cross-validation (LOOCV). For each held-out patient, the volume-adjusted score map was recalculated using the remaining 55 patients, with eligibility determined by lesion coverage in the training set. A training-set peak was selected, and the peak-plus-volume model was fitted using those 55 patients. Log-volume standardization was also estimated within the training set. This model then generated the held-out patient’s probability. Discrimination was summarized by the area under the receiver operating characteristic curve (AUC) across all 56 out-of-fold predictions.

Significance of the model’s overall discrimination was assessed by 2,000 random permutations of the outcome labels, preserving the number of positive outcomes. Every permutation repeated the entire LOOCV pipeline, including training-set map estimation, peak selection and model fitting. The one-sided permutation P value was the proportion of permuted AUCs at least as large as the observed AUC, calculated with a plus-one correction.

### Validation in separate cohorts

For validation, the model was estimated using all 56 development patients. The selected peak, regression coefficients, intercept and log-volume standardization parameters were then held fixed when predicting outcomes in the two validation cohorts. Discrimination was evaluated in the pooled sample of 19 patients; cohort-specific performance was reported as a descriptive breakdown. No model refitting or threshold recalibration was performed in the validation cohorts.

A 95% confidence interval for the pooled AUC was obtained from 50,000 percentile bootstrap samples, resampling patients within each cohort-by-outcome stratum while holding the model fixed. Statistical significance was assessed using an exact permutation test with fixed predictions and outcome labels permuted within each cohort while preserving its positive count. All 25,740 allowable label arrangements were enumerated. The two-sided P value was twice the smaller inclusive tail probability, capped at one.

The volume-only reference was fitted in the development cohort using the same Firth/FLIC approach and applied without refitting to the validation patients. The AUCs of the two fixed models were compared in the pooled validation sample using a two-sided paired DeLong test.[38] This comparison evaluated the difference in discrimination between the models, separately from the exact permutation test of the combined model’s overall discrimination.

For threshold-dependent validation summaries, a probability cut-off maximizing Youden’s index was derived from the development-cohort LOOCV predictions and applied unchanged to validation patients (0.2977). Sensitivity and specificity were reported with exact binomial 95% confidence intervals. The Brier score summarized overall probability prediction error.

### Overlap with a structural map of consciousness recovery

The supplementary exploratory analysis used Warren and colleagues’ publicly released optimal structural connectivity map (DBS_DoC_OptimalStructConn; https://osf.io/bjah5/).[11] Its image dimensions and affine transformation matched the 0.5-mm isotropic lesion grid, allowing direct sampling at lesion voxel centres without additional spatial interpolation for this comparison.

For each development patient, map-weighted lesion burden was the sum of positive map intensities within the entire lesion, with non-positive background set to zero. This score retained map intensity and lesion extent; multiplication by 0.125 mm³ gives the corresponding weighted integral. Groups were compared using a two-sided Mann–Whitney U test with tie and continuity corrections. P values were nominal, without lesion-volume adjustment.

### Statistical analysis

Lesion volumes were summarized as medians and interquartile ranges and compared between outcome groups using a two-sided Mann–Whitney U test. One-sided tests were used for the positive voxel-wise association and the development-model permutation test; pooled validation inference was two-sided. Statistical significance was defined as P < 0.05, with FWER-corrected P values used for voxel-wise localization. Analyses were implemented in Python using NumPy, SciPy, pandas, NiBabel and statsmodels, with custom implementations of Firth/FLIC fitting and resampling procedures.

### Code availability

The analysis code and fixed model parameters are supplied as Supplementary Software 1 and will be made publicly available on GitHub upon publication.

## Results

### Participants and lesion characteristics

The development cohort included 56 patients, of whom 14 (25.0%) had somnolence and 42 did not. Lesions were originally left-sided in 40 patients and right-sided in 16. Median lesion volume was 63.6 mm³ (interquartile range [IQR] 43.3–87.4). Patients with somnolence had larger lesions than those without somnolence: 91.4 mm³ (IQR 68.1–128.2) versus 57.4 mm³ (IQR 39.8–80.7; two-sided Mann–Whitney U test, P = 0.0017). Representative perioperative video recordings documented preserved interaction before lesioning, a subsequent reduction in arousal and recovery.

The validation cohorts comprised 13 patients with movement disorders treated at FJK Tokyo Hospital and six patients with epilepsy treated at Tokyo Women’s Medical University. Somnolence was present in seven of 13 (53.8%) and four of six (66.7%) patients, respectively, giving 11 of 19 (57.9%) patients in the pooled validation sample. Median lesion volumes were 153.4 mm³ (IQR 125.8–224.4) in the FJK cohort and 120.3 mm³ (IQR 82.6–181.1) in the epilepsy cohort (Table 1).

**Table 1.** Clinical and lesion characteristics of the study cohorts.

| Characteristic | Development cohort |  |  | Validation cohorts |  |
|---|---|---|---|---|---|
|  | All patients<br>(n = 56) | Somnolence<br>(n = 14) | No somnolence<br>(n = 42) | FJK movement disorders<br>(n = 13) | TWMU epilepsy<br>(n = 6) |
| Procedure period | 2017–2022 | 2017–2022 | 2017–2022 |  |  |
| Primary surgical indication | Movement disorder | Movement disorder | Movement disorder | Movement disorder | Epilepsy |
| Parkinson disease, n (%) |  |  |  |  | 0 (0.0) |
| Dystonia, n (%) |  |  |  |  | 0 (0.0) |
| Other movement disorders, n (%) |  |  |  |  | 0 (0.0) |
| Epilepsy, n (%) | 0 (0.0) | 0 (0.0) | 0 (0.0) | 0 (0.0) | 6 (100.0) |
| Surgical side, left, n (%) | 40 (71.4) | 7 (50.0) | 33 (78.6) | 0 (0.0) | 3 (50.0) |
| Surgical side, right, n (%) | 16 (28.6) | 7 (50.0) | 9 (21.4) | 13 (100.0) | 3 (50.0) |
| Lesion volume, mm<sup>3</sup><br>median (IQR) | 63.6<br>(43.3–87.4) | 91.4<br>(68.1–128.2) | 57.4<br>(39.8–80.7) | 153.4<br>(125.8–224.4) | 120.3<br>(82.6–181.1) |
| Somnolence, n (%) | 14 (25.0) | 14 (100.0) | 0 (0.0) | 7 (53.8) | 4 (66.7) |

Values are n (%) or median (IQR). IQR, interquartile range; TWMU, Tokyo Women’s Medical University. Diagnosis refers to the primary indication for PTT. Surgical side is the original side before left alignment.
Development-cohort lesion-volume comparison between patients with and without somnolence: two-sided Mann–Whitney U test, P = 0.0017.

### Volume-adjusted probabilistic lesion mapping

A total of 2,074 voxels met the requirement for lesion coverage in at least five patients. Volume-adjusted mapping identified two voxels with FWER-corrected P < 0.05. The largest positive score was located at MNI coordinates (−6.0, −10.5, −4.0) mm (Z = 4.33; FWER-corrected P = 0.0111). This voxel was included in the lesions of four of 14 patients with somnolence and one of 42 patients without somnolence. Atlas comparison placed the peak dorsal to the lateral hypothalamus, 3.52 mm from its nearest atlas voxel centre (Fig. 1; Supplementary Fig. S1; Table 2).

**Figure 1.**
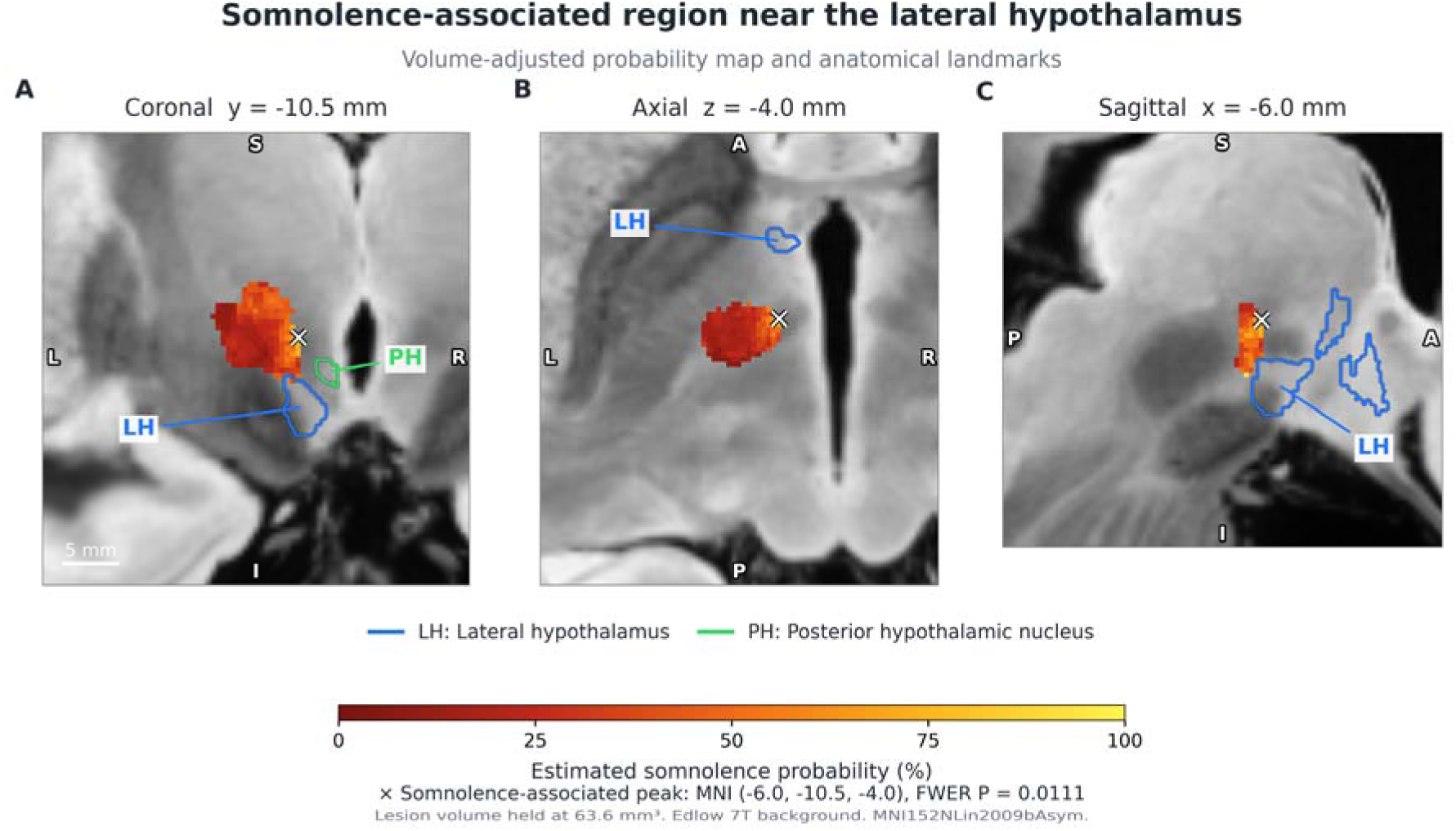
Anatomical location of somnolence-associated lesions. Continuous colours show fitted somnolence probability after adjustment for log-transformed lesion volume, evaluated at the development median volume of 63.6 mm³. The cross marks the volume-adjusted statistical association peak used in the prediction model, MNI (−6.0, −10.5, −4.0) mm (FWER-corrected P = 0.0111). Blue, lateral hypothalamus; green, posterior hypothalamic nucleus. Colours represent fitted probability, with anatomical contours as reference labels. Coronal, axial and sagittal sections pass through the statistical peak in MNI152NLin2009bAsym space. The anatomical background is the Edlow 7T MRI dataset (Edlow et al., Scientific Data 2019;6:244, doi:10.1038/s41597-019-0254-8); the hypothalamic atlas is from Neudorfer et al. (Scientific Data 2020;7:305, doi:10.1038/s41597-020-00644-6).

**Table 2.** Anatomical localization of somnolence-associated lesions.

| Location definition | MNI coordinates<br>(x, y, z), mm | Z | FWER-corrected<br>P | Lesion involvement<br>somnolence, n/N | Lesion involvement<br>no somnolence, n/N | Adjusted fitted<br>probability (%) |
|---|---|---|---|---|---|---|
| Peak statistical association | −6.0, −10.5, −4.0 | 4.33 | 0.0111 | 4/14 | 1/42 | 95.3 |
| Highest fitted probability | −6.0, −12.0, −9.5 | 4.15 | 0.0230 | 4/14 | 1/42 | 96.8 |
| Unadjusted Fisher exact peak | −7.5, −11.0, −3.5 | — | 0.00895 | 11/14 | 7/42 | — |

Development cohort: 56 patients, including 14 with and 42 without somnolence. The search volume comprised 2,074 voxels lesioned in at least five patients. Coordinates are shown in left-aligned MNI152NLin2009bAsym space.
FWER, family-wise error. Adjusted P values are one-sided FWER values from 20,000 parametric bootstrap replicates for volume-adjusted scores and 20,000 label permutations for Fisher tests. Z and P at the maximum-probability location are its volume-adjusted score-test values.
Fitted probabilities assume involvement of the specified voxel at the development-cohort median lesion volume of 63.6 mm<sup>3</sup>. The maximum volume-adjusted score lies dorsal to the lateral hypothalamus, 3.52 mm from its nearest atlas voxel centre. —, value not displayed for that item.
In the pooled validation cohort, the statistical association peak used for prediction was included in lesions of six of 11 patients with somnolence and one of eight without somnolence.

On the continuous probability map, the highest fitted probability was 96.8% at (−6.0, −12.0, −9.5) mm, evaluated at the development-cohort median lesion volume of 63.6 mm³. This location also showed a positive volume-adjusted association with somnolence (Z = 4.15; FWER-corrected P = 0.0230). The fitted probability at the maximum-score voxel was 95.3% at the same reference volume. The maximum-score voxel was the location used in the peak-plus-volume prediction model. The spatial distribution of the probability map is illustrated in Fig. 2 and Supplementary Figs. S2–S5.

**Figure 2.**
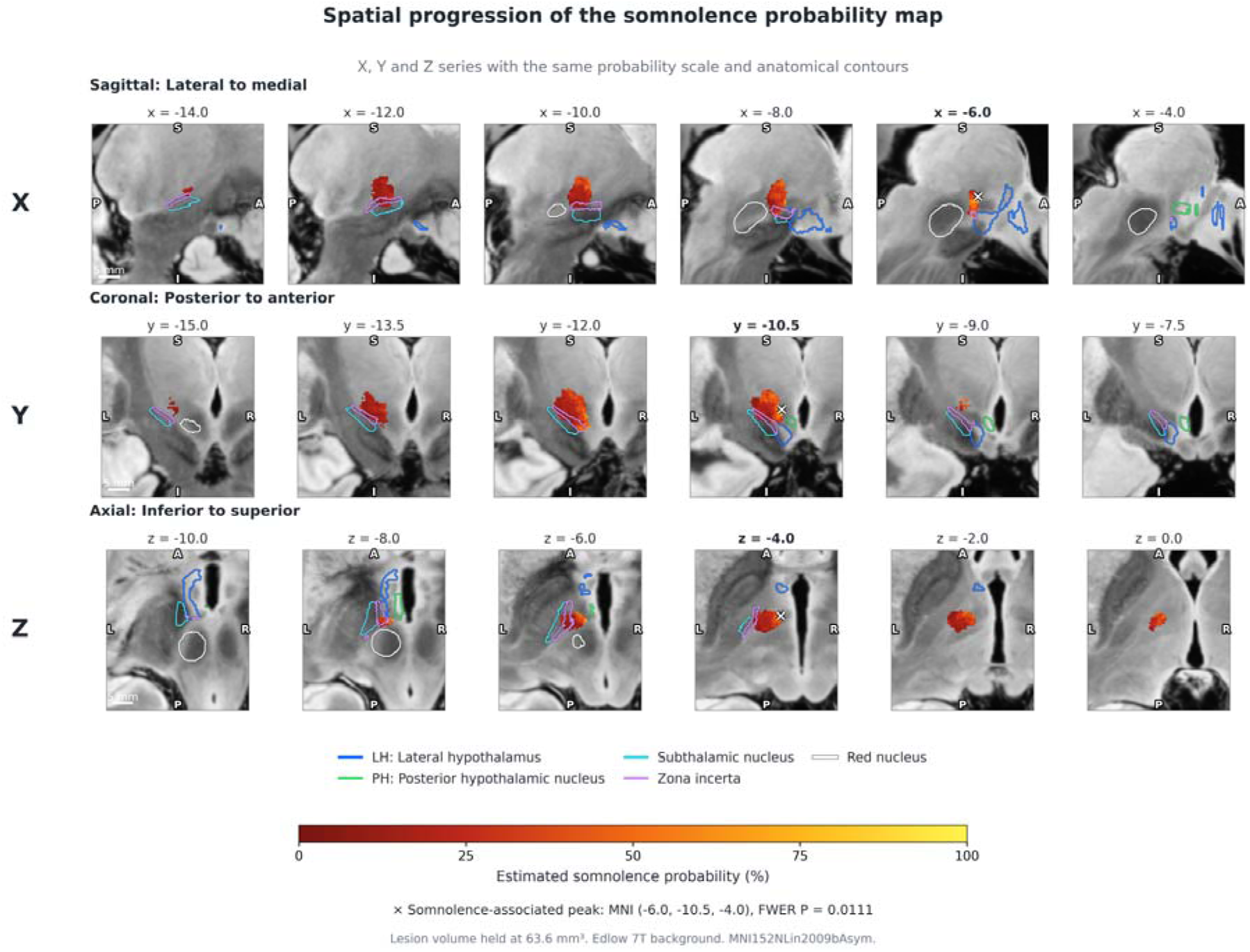
Spatial extent across serial MNI sections. Serial sagittal, coronal and axial sections show the same probability map and fixed 0–100% colour scale as Figure 1. Cyan, subthalamic nucleus; purple, zona incerta; white, red nucleus. The cross appears only in a plane intersecting the statistical peak. Slice coordinates and orientations are printed in each panel.

### Supporting and sensitivity analyses

Unadjusted Fisher mapping peaked at (−7.5, −11.0, −3.5) mm (permutation FWER-corrected P = 0.00895). At this voxel, lesions were present in 11 of 14 patients with somnolence and seven of 42 patients without somnolence. The two-sided volume-adjusted analysis retained the same maximum-score coordinate (FWER-corrected P = 0.0111). After excluding the two cases with atypical lesion locations, the remaining 54 patients included 14 with somnolence. The volume-adjusted maximum remained at (−6.0, −10.5, −4.0) mm (Z = 4.20; one-sided FWER-corrected P = 0.0177).

### Internal predictive performance

The peak-plus-volume model achieved an LOOCV AUC of 0.799 (Fig. 3). The complete-pipeline permutation test yielded a one-sided P value of 0.0020 (Fig. 4). The out-of-fold Brier score was 0.148. The volume-only reference model had an LOOCV AUC of 0.745 and a Brier score of 0.160 (Table 3).

**Figure 3.**
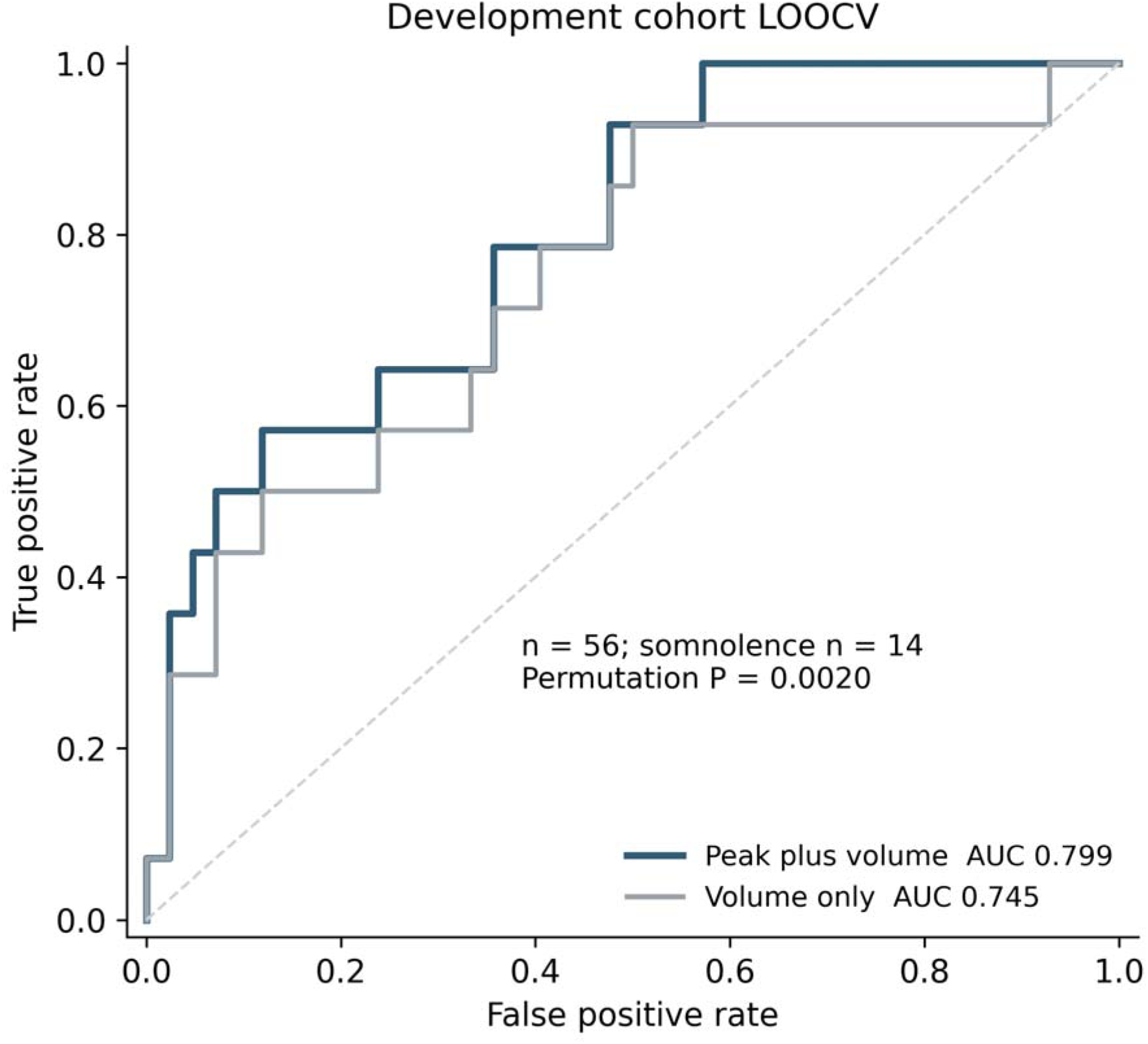
Internal cross-validation. Empirical ROC curves from out-of-fold predictions in 56 patients. For each held-out patient, the volume-adjusted map, peak selection, log-volume standardization and model fitting used only the remaining 55 patients. The peak-plus-volume model achieved an AUC of 0.799; the volume-only reference achieved 0.745. P = 0.0020 tests overall discrimination of the combined model by a one-sided complete-pipeline label permutation test; it does not test the AUC difference between models.

**Figure 4.**
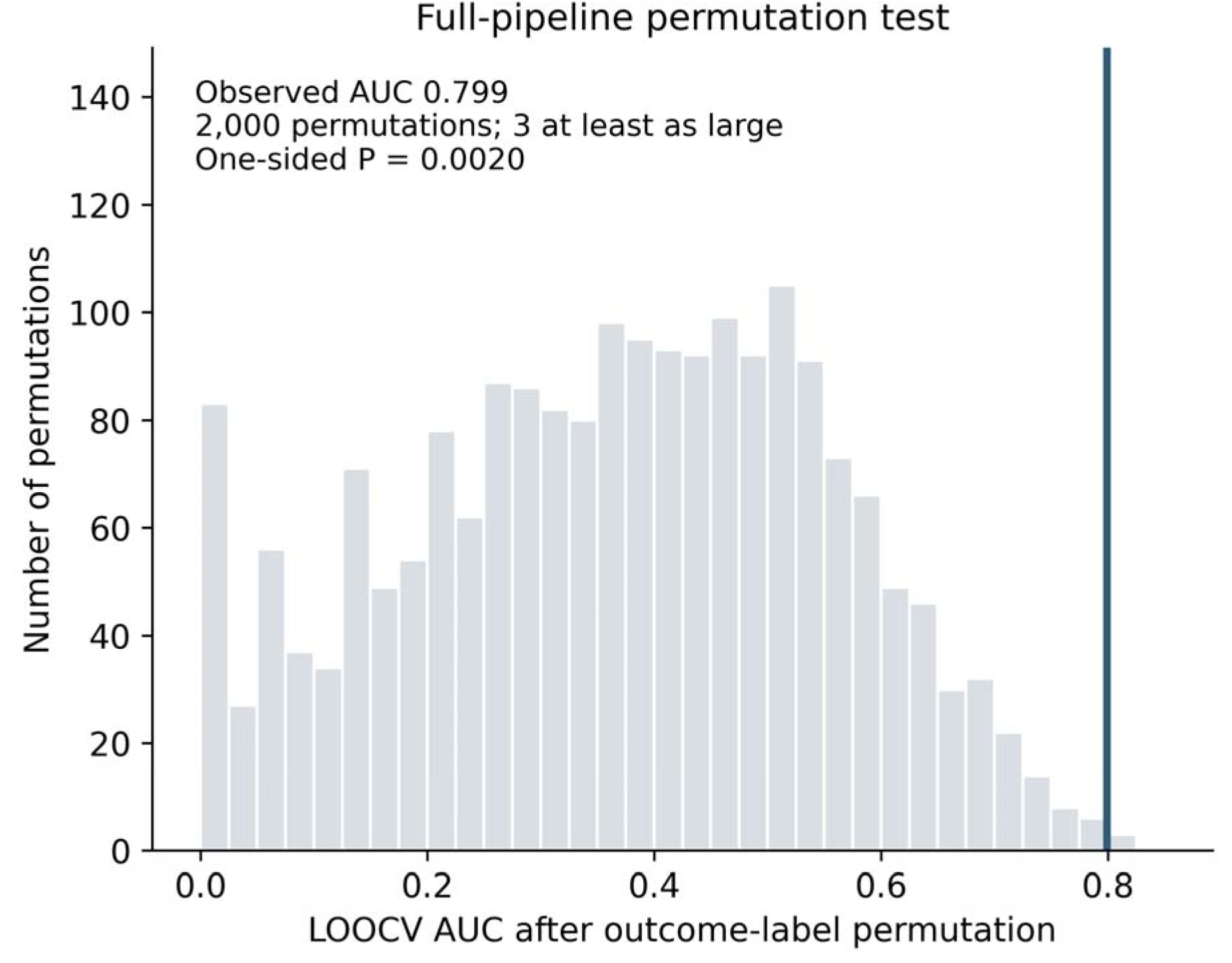
Permutation distribution of internal discrimination. Distribution of 2,000 LOOCV AUCs after permutation of the outcome labels while preserving the positive count. Each permutation repeated mapping, peak selection, log-volume standardization and model fitting within LOOCV. The blue line marks the observed AUC of 0.799. Three permutations reached or exceeded it; the plus-one P value was (3 + 1)/(2,000 + 1) = 0.0020.

**Table 3.**
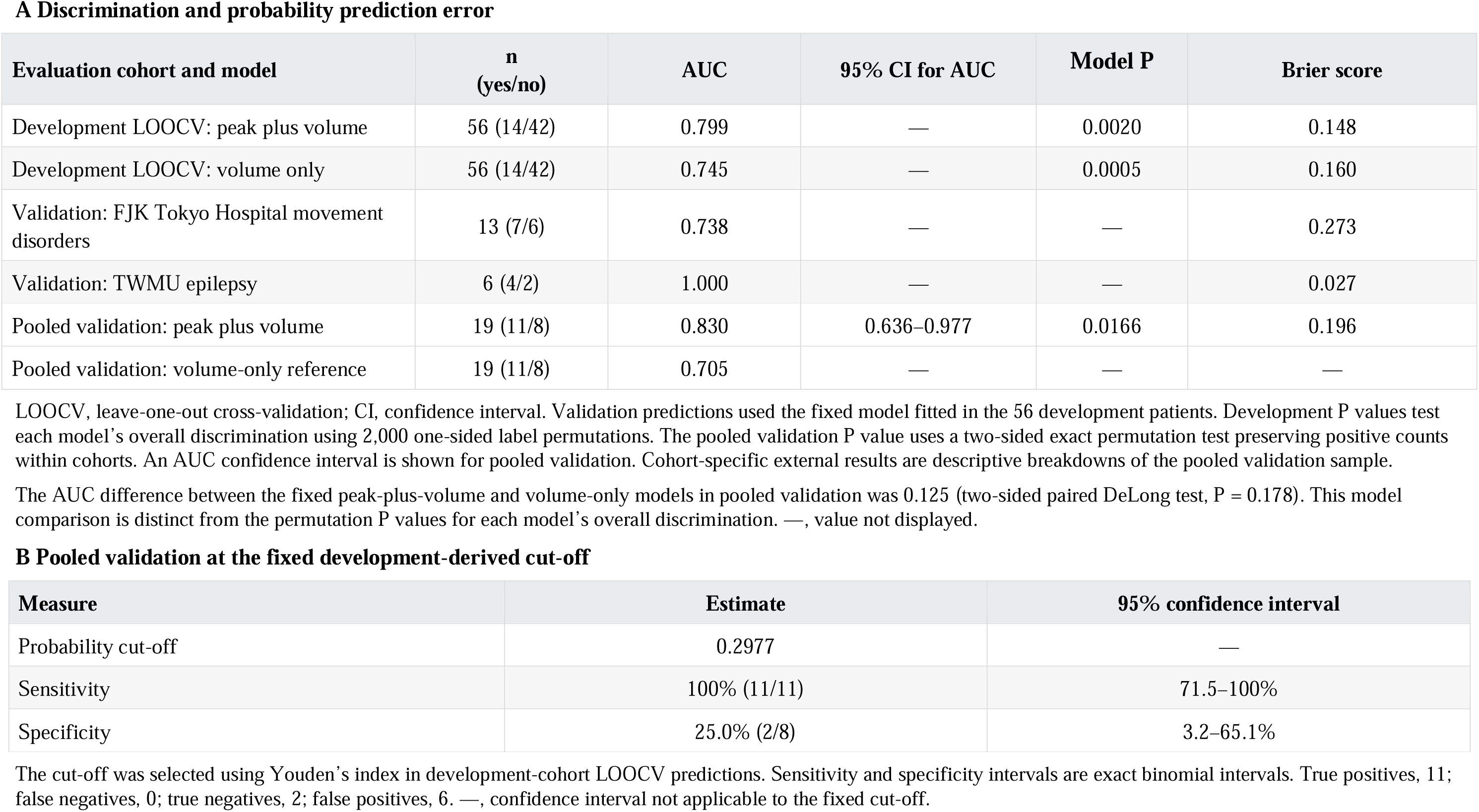
Predictive performance in development and validation cohorts.

| **A Discrimination and probability prediction error** |  |  |  |  |  |
| **Evaluation cohort and model** | **n<br>(yes/no)** | **AUC** | **95% CI for AUC** | **Model P** | **Brier score** |
|---|---|---|---|---|---|
| Development LOOCV: peak plus volume | 56 (14/42) | 0.799 | — | 0.0020 | 0.148 |
| Development LOOCV: volume only | 56 (14/42) | 0.745 | — | 0.0005 | 0.160 |
| Validation: FJK Tokyo Hospital movement disorders | 13 (7/6) | 0.738 | — | — | 0.273 |
| Validation: TWMU epilepsy | 6 (4/2) | 1.000 | — | — | 0.027 |
| Pooled validation: peak plus volume | 19 (11/8) | 0.830 | 0.636–0.977 | 0.0166 | 0.196 |
| Pooled validation: volume-only reference | 19 (11/8) | 0.705 | — | — | — |
| **B Pooled validation at the fixed development-derived cut-off** |  |  |  |  |  |
| **Measure** | **Estimate** | **95% confidence interval** |  |  |  |
| Probability cut-off | 0.2977 | — |  |  |  |
| Sensitivity | 100% (11/11) | 71.5–100% |  |  |  |
| Specificity | 25.0% (2/8) | 3.2–65.1% |  |  |  |

LOOCV, leave-one-out cross-validation; CI, confidence interval. Validation predictions used the fixed model fitted in the 56 development patients. Development P values test each model’s overall discrimination using 2,000 one-sided label permutations. The pooled validation P value uses a two-sided exact permutation test preserving positive counts within cohorts. An AUC confidence interval is shown for pooled validation. Cohort-specific external results are descriptive breakdowns of the pooled validation sample.
The AUC difference between the fixed peak-plus-volume and volume-only models in pooled validation was 0.125 (two-sided paired DeLong test, P = 0.178). This model comparison is distinct from the permutation P values for each model’s overall discrimination. —, value not displayed.
The cut-off was selected using Youden’s index in development-cohort LOOCV predictions. Sensitivity and specificity intervals are exact binomial intervals. True positives, 11; false negatives, 0; true negatives, 2; false positives, 6. —, confidence interval not applicable to the fixed cut-off.

### Validation in separate cohorts

Applied without refitting, the development model achieved an AUC of 0.830 in the pooled validation sample of 19 patients (95% bootstrap confidence interval 0.636–0.977; Fig. 5a). The two-sided cohort-stratified exact permutation P value was 0.0166. Cohort-specific AUCs, reported as descriptive breakdowns, were 0.738 in the FJK movement-disorder cohort and 1.000 in the six-patient epilepsy cohort (Fig. 5b).

**Figure 5.**
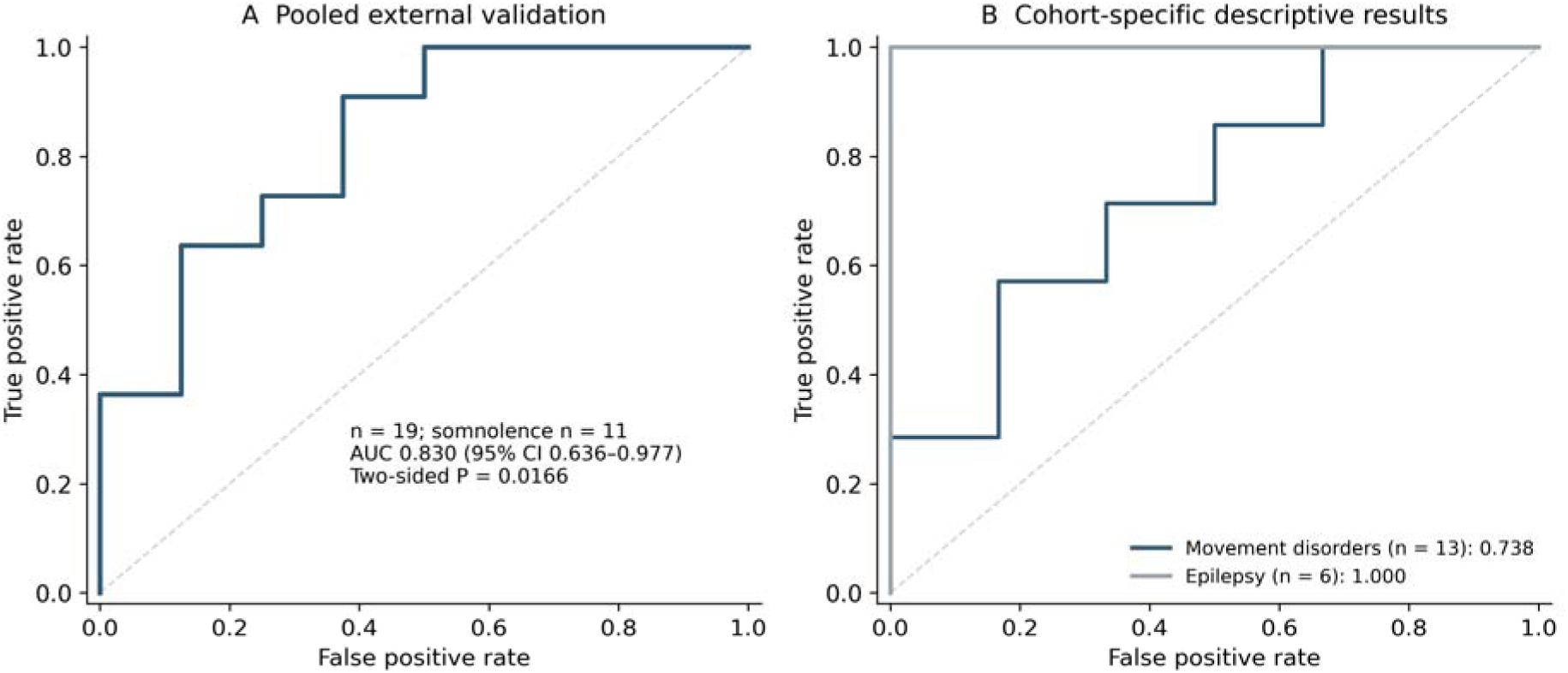
External validation of the fixed model. A, ROC curve for the model fitted in all 56 development patients and applied without refitting to 19 validation patients (AUC 0.830; 95% CI 0.636–0.977). The confidence interval used 50,000 cohort-by-outcome stratified bootstrap resamples. The two-sided P value of 0.0166 used all 25,740 within-cohort label allocations and tests overall discrimination of the combined model. B, Cohort-specific ROC curves are descriptive breakdowns of the same fixed predictions. The volume-only reference had a pooled validation AUC of 0.705; the paired model comparison is reported in Table 3.

In the pooled validation sample, the development-defined association peak was included in lesions of six of 11 patients with somnolence and one of eight without somnolence. The fixed volume-only reference model had an AUC of 0.705, compared with 0.830 for the peak-plus-volume model (AUC difference 0.125; two-sided paired DeLong test, P = 0.178; Table 3).

Individual predicted probabilities in the development and validation cohorts are shown in Fig. 6. At the fixed development-derived probability cut-off of 0.2977, all 11 patients with somnolence were classified as positive, and two of eight patients without somnolence were classified as negative. Sensitivity was 100% (11/11; exact 95% confidence interval 71.5–100%) and specificity was 25.0% (2/8; exact 95% confidence interval 3.2–65.1%; Fig. 7). The pooled validation Brier score was 0.196 (Table 3).

**Figure 6.**
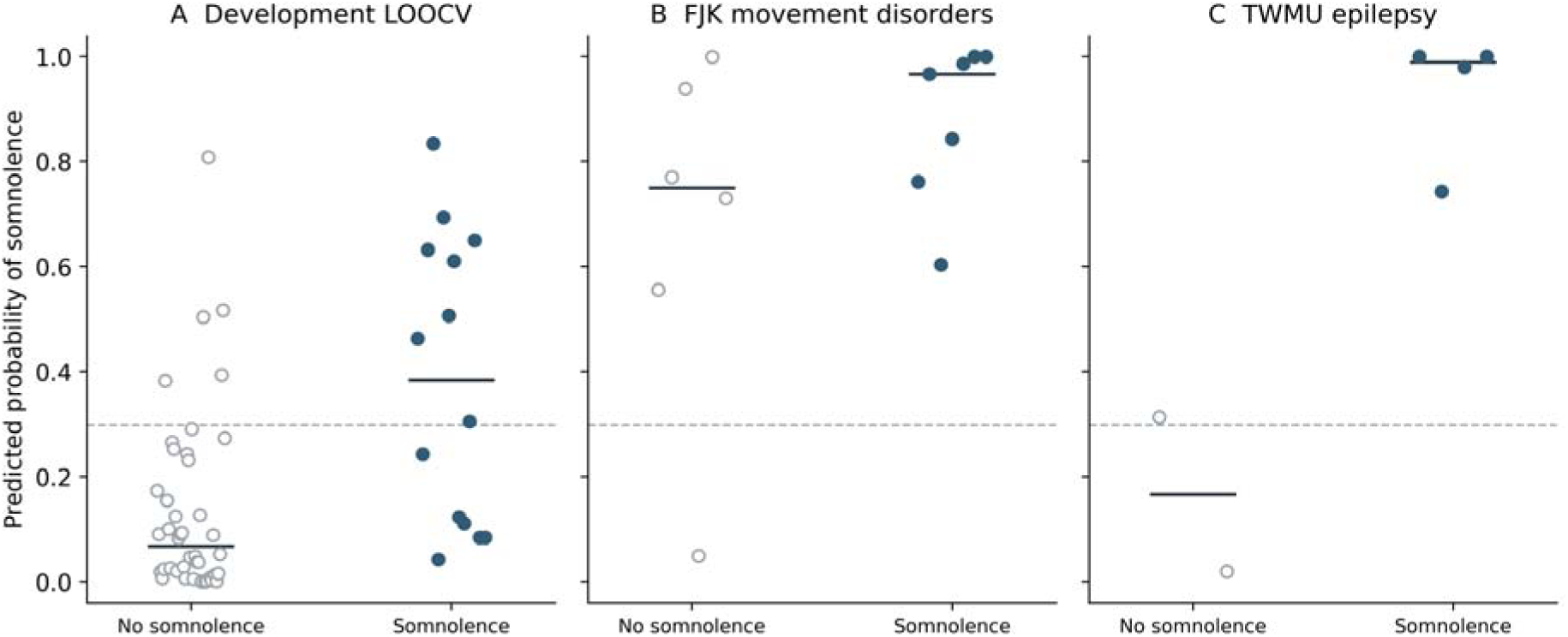
Patient-level predicted probabilities. Each point represents one patient; short black lines indicate group medians. A, Development-cohort out-of-fold probabilities. B and C, Fixed-model predictions in the validation cohorts. Filled blue points indicate somnolence; open grey points indicate no somnolence. Horizontal offsets prevent overlap. The dashed line is the fixed development-derived threshold of 0.2977.

**Figure 7.**
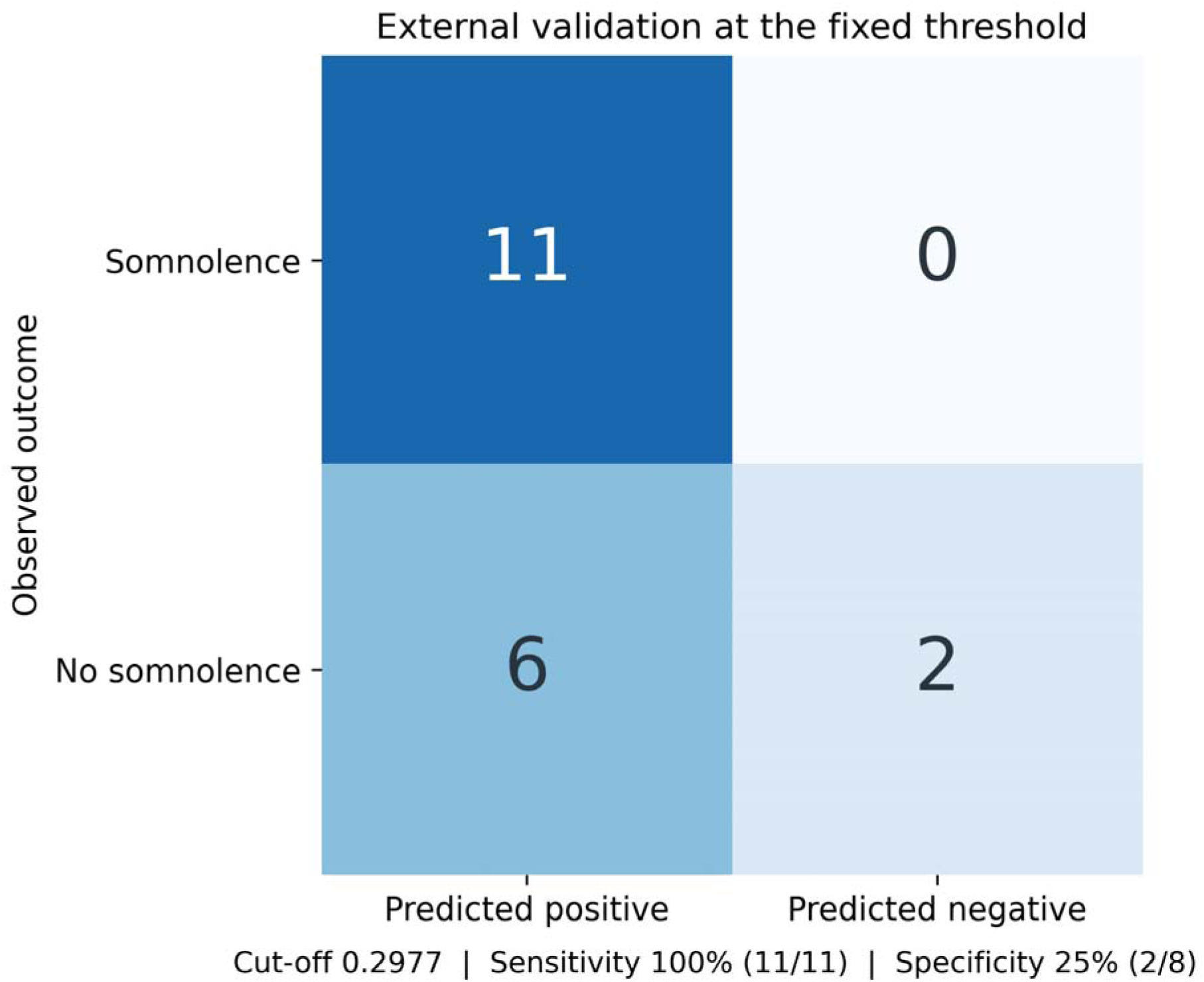
Classification at the development-derived threshold. Confusion matrix for the 19 validation patients at the unchanged probability cut-off of 0.2977. Rows represent observed outcomes and columns represent model classifications. There were 11 true positives, no false negatives, two true negatives and six false positives. Sensitivity was 100% (exact 95% CI 71.5–100%); specificity was 25.0% (3.2–65.1%).

### Overlap with a structural map of consciousness recovery

In a supplementary anatomical analysis, lesions from the 56 development patients were compared with the published structural connectivity map associated with recovery of consciousness after DBS reported by Warren and colleagues.[11] Map-weighted lesion burden was quantified as the sum of positive map intensities within each entire lesion, integrating map intensity with the amount of map territory encompassed by the lesion. This score was higher in patients with somnolence than in those without somnolence (median 167.23 [IQR 49.52–260.48] versus 56.38 [IQR 8.85–81.94]; two-sided Mann–Whitney U test, P = 0.0299). The supplementary comparison retained the contribution of lesion extent and was performed without adjustment for total lesion volume.

## Discussion

Focal PTT lesions identified a subthalamic territory associated with transient impairment of wakefulness. The spatial association persisted after adjustment for lesion volume and multiple comparisons, and a model combining involvement of the association peak with lesion volume discriminated somnolence in 19 additional patients treated by other surgeons. These findings locate a vulnerability of human wakefulness within a compact region traversed during surgery for motor disorders. Comparison of nearby lesions with different outcomes focuses the mechanistic question on the distribution of injured tissue and connections within a shared surgical territory. Supplementary analysis showed greater map-weighted lesion burden within Warren and colleagues’ structural map of stimulation-related consciousness recovery in patients with somnolence.[11]

The association peak at MNI (−6.0, −10.5, −4.0) was dorsal to the lateral hypothalamus, with a Euclidean distance of 3.52 mm to the nearest voxel centre in the hypothalamic atlas.[12] Its interpretation requires the surrounding fibre anatomy. The lenticular fasciculus courses through Forel’s field H2 and joins the ansa lenticularis to form the thalamic fasciculus, or field H1; the zona incerta lies between H1 and H2.[3] These relationships define a compact neighbourhood containing pallidal efferents, incertal tissue and the hypothalamic border. The peak localizes the strongest statistical association within the sampled lesions. Assigning it to a particular fascicle or neuronal population requires greater anatomical specificity than this coordinate and atlas distance provide.

Our observations extend an established clinical phenomenon into an anatomical comparison. Transient somnolence after PTT has been described by Gallay, Godinho and our group, and postoperative polysomnography has documented changes in sleep and waking EEG.[4–7] In the present cohort, surgery was performed while patients were awake and without systemic sedation. Representative recordings captured interaction before lesioning, reduced responsiveness afterwards and subsequent recovery. This temporal sequence supports an acute effect of the intervention on wakefulness. Comparison with patients undergoing the same procedure without somnolence identifies spatial variation within a shared operative region, providing information beyond the occurrence of sleepiness after hypothalamic-region injury.

Von Economo’s observations in encephalitis lethargica implicated the posterior hypothalamic–rostral midbrain region in sleep–wake regulation.[13] Ranson’s primate lesions and Nauta’s rat transections subsequently examined this localization experimentally.[14,15] Nauta distinguished sustained waking without external stimulation from responsiveness to stimulation and implicated fibres traversing the lateral hypothalamus. Moruzzi and Magoun’s stimulation experiments then showed widespread EEG activation from brainstem, dorsal hypothalamic and subthalamic sites.[16] Together, these studies make the present border localization biologically relevant while leaving a central question unresolved: whether regional injury impairs local neurons, the transmission of ascending activity, or both. Thermal lesions can affect each component.

Subsequent experiments identified routes by which brainstem activity reaches the forebrain. Fuller and colleagues found profoundly different effects of extensive thalamic lesions and lesions of the parabrachial–precoeruleus complex or basal forebrain in rats.[17] Qiu and colleagues showed that activation of parabrachial neurons projecting to the lateral/posterior hypothalamus or preoptic–basal forebrain region promoted sustained waking, whereas activation of the thalamus-projecting population did not produce a comparable sustained effect.[18] The hypothalamic region therefore participates in an extrathalamic route for behavioural arousal as well as in local state regulation. Our dorsal border localization is consistent with injury reducing the input to, or output from, wake-promoting populations while leaving much of the distributed arousal system intact.

The lateral hypothalamus also contributes to the stability of waking. Human orexin neurons occupy the posterior perifornical and lateral hypothalamic region and are markedly depleted in narcolepsy.[8,9] Orexin-deficient mice have fragmented behavioural states despite near-normal total sleep–wake amounts and can sustain waking in a novel environment.[19] Extensive orexin projections to arousal-related structures,[20] together with awakening after orexin activation and sleep after acute silencing,[21,22] provide mechanisms through which disturbed hypothalamic function could impair continuity of wakefulness. This region additionally contains wake-promoting GABAergic neurons, including a population that inhibits sleep-promoting preoptic neurons.[23,24] Repeated postoperative sleep episodes may reflect impaired maintenance of waking; the profound hyporesponsiveness seen in some patients indicates that the clinical spectrum can extend beyond this manifestation.

The neighbouring zona incerta prevents a simple equation between hypothalamic proximity and loss of wake-promoting neurons. Liu and colleagues identified ventral incertal Lhx6-positive GABAergic neurons that inhibit wake-active orexin and GABAergic hypothalamic cells; activating these neurons promoted sleep, whereas inhibiting them reduced sleep.[25] Selective ablation of MCH neurons in another mouse study increased wakefulness and reduced NREM sleep.[26] Loss of these sleep-promoting populations alone would therefore not straightforwardly explain postoperative somnolence. The anatomical neighbourhood contains populations with different effects on vigilance, and regional thermal injury need not reproduce selective cellular inhibition. The observed direction of impairment makes involvement of wake-supporting cells or connections a more plausible hypothesis than an undifferentiated effect of damaging any sleep-related tissue nearby.

Interruption of pallidal output remains an alternative or contributing explanation. PTT targets pallidothalamic fibres.[3] Earlier clinical reports proposed pallidomesencephalic disturbance as a mechanism of somnolence.[4,5] Single-axon tracing in primates demonstrated extensively collateralized GPi projections to thalamic nuclei and the pedunculopontine nucleus, with pallidofugal axons traversing Forel’s field H.[27] Pallidal and arousal-related mechanisms may consequently interact within this region. All development patients underwent PTT, yet only a subset developed somnolence, and its spatial association persisted after adjustment for total lesion volume. This argues for a contribution from the distribution of injury within the operative territory. The comparison cannot distinguish differential interruption of pallidal projections from additional involvement of neighbouring arousal pathways, and the two mechanisms may coexist.

Human studies place this focal vulnerability within a wider ascending system. Fischer and colleagues compared 12 coma-causing brainstem lesions with 24 controls to identify a rostral dorsolateral pontine region, while all four patients with coma or stupor among 33 thalamic strokes in Hindman and colleagues’ study had brainstem extension.[2,28] Edlow and colleagues reconstructed connections among brainstem, hypothalamic, thalamic and basal forebrain nodes in three postmortem brains, complemented by in vivo imaging.[1] Their ventral tegmental and medial forebrain bundle pathways provide an anatomical framework for considering connecting tissue near our hotspot. Our transient postoperative phenotype differs from coma, but both settings illustrate how strategically situated injury can impair arousal within a distributed system.

The supplementary comparison with the structural map reported by Warren and colleagues provides an anatomical context for our lesion findings. Among 40 patients receiving DBS for disorders of consciousness, structural analysis of 28 successfully registered cases identified improvement-associated connections including a bundle resembling the ventral tegmental tract; ten-fold cross-validation supported the structural association (P = 0.03).[11] In our cohort, the summed positive map values within lesions were greater with somnolence (P = 0.0299). This measure integrates the intensity of the published map with the amount of its territory encompassed by each lesion. Its rationale is that symptoms may depend on the cumulative amount of functionally relevant tissue or connections affected. This supplementary association therefore includes lesion extent; an effect independent of total lesion volume or specificity relative to other maps was not established. Central thalamic stimulation has improved responsiveness in a patient minimally conscious for six years and restored arousal in anaesthetized macaques.[29,30] Stimulation studies examine the recruitment of circuitry to increase arousal, whereas our intervention perturbs an established waking state. The anatomical correspondence motivates the hypothesis that pathways implicated in consciousness recovery also contribute to maintaining wakefulness.

Recovery despite a persisting lesion can be understood by separating structural loss from the evolving function of spared circuitry. Denoyer and colleagues found marked acute hypersomnia after posterior hypothalamic neuronal lesions in cats, with sleep measures returning to control levels by day three; combined hypothalamic and mesencephalic lesions initially produced coma-like behaviour followed by recovery.[31] Blanco-Centurion and colleagues found preserved daily wake at three weeks despite substantial combined lesions of basal forebrain cholinergic, tuberomammillary histaminergic and locus coeruleus noradrenergic populations.[32] These experiments differ from acute human PTT, but demonstrate that the consequences of injury depend on time and on the remaining system. Spared ipsilateral pathways, contralateral connections and other populations may support recovery. A unilateral lesion alone does not establish which of these processes produced the transient clinical course.

Reversible peri-lesional dysfunction may contribute concurrently. Godinho and colleagues described increased oedema in two of four patients with transient somnolence,[5] and sleepiness after a small hypothalamic infarction has resolved by the following day.[10] MRI within 24 hours captures tissue during this changing acute phase. Functional disturbance may extend beyond the tissue ultimately destroyed, and its resolution can restore adequate activity in surviving pathways. The transient symptoms therefore remain compatible with a contribution of the injured territory to normal wakefulness. Their resolution motivates consideration of functional reserve and reversible disturbance without requiring that compensation arise exclusively from the opposite hemisphere.

The predictive results support relevance beyond the original surgeon’s cohort. With mapping, peak selection and model fitting repeated within each training set, the peak-plus-volume model achieved an LOOCV AUC of 0.799. Without refitting, its AUC was 0.830 in the pooled movement-disorder and epilepsy validation cohorts (95% confidence interval 0.636–0.977; two-sided cohort-stratified exact permutation P = 0.0166). These results describe the combined model’s discrimination; an increment in external AUC over the volume-only reference was not statistically established. Location and extent have complementary biological meanings: a lesion can involve a strategically situated connection, a larger amount of relevant tissue, or both. The model does not imply that destruction of a single voxel is sufficient to cause somnolence or establish a biological injury threshold. The fixed development-derived cut-off yielded 100% sensitivity and 25% specificity externally, so clinical use requires further evaluation of calibration and decision thresholds.

These observations add arousal-related anatomy to surgical planning as PTT expands to staged bilateral treatment. The FDA’s July 2025 indication expansion permits contralateral PTT at least six months after the initial procedure in eligible patients with Parkinson’s disease.[33] In a prospective multicentre study, ten of 40 bilaterally treated patients had persistent moderate or severe treatment-related adverse events at 12 months, predominantly affecting speech, gait and balance.[34] Our results provide a spatial reference for considering wakefulness alongside motor outcomes. They do not determine the effect of any particular bilateral lesion configuration, which depends on the tissue and connections involved.

The retrospective analysis included patients with sufficient documentation to determine somnolence; it cannot quantify symptoms in excluded patients. The binary outcome combines impaired wake maintenance and reduced responsiveness without EEG-defined state or standardized severity and duration measurements. Disease and medication effects were not included in the prediction model. Acute MRI segmentation and atlas registration limit anatomical precision, and only five development patients covered the association peak. Validation in 19 patients yields a wide AUC confidence interval. The Warren comparison uses a group-level structural map rather than patient-specific axonal measurements. Evidence from animal experiments identifies candidate mechanisms but does not establish which human cells or fibres were injured in this cohort. These constraints define the level of interpretation: the study localizes a clinically relevant surgical territory, while its cellular composition, affected projections and recovery physiology remain to be resolved.

The principal contribution is a human lesion-based localization of vulnerability in maintaining wakefulness, with discrimination of outcomes in additional surgical cohorts. The findings connect disruption of a distributed arousal function with the location and extent of focal injury within a compact subthalamic surgical territory.

## Data Availability

De-identified data supporting the findings of this study are available from the corresponding author upon reasonable request, subject to institutional approval and applicable data-sharing restrictions.

---

**Supplementary Figure S1.**
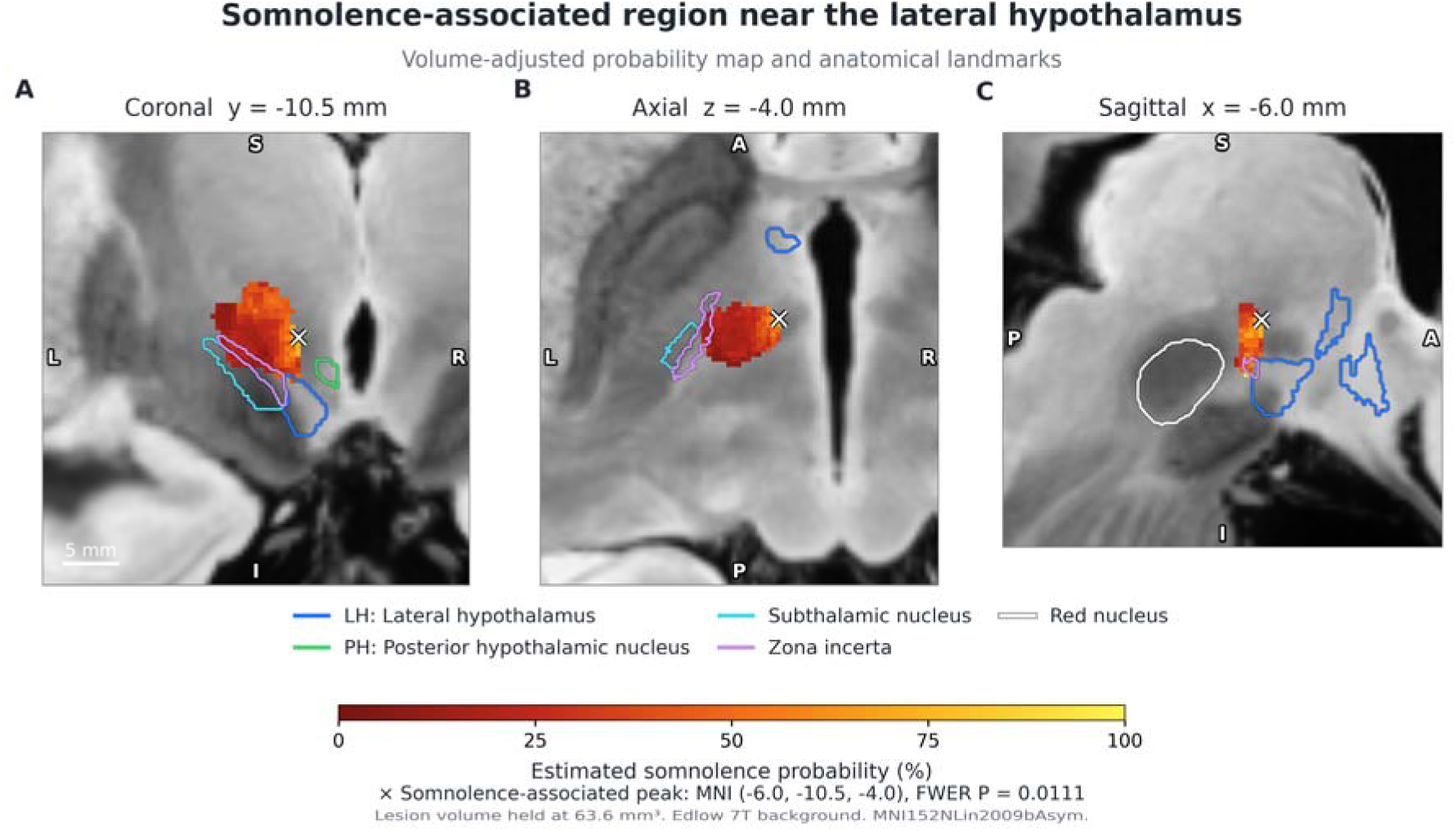
Three-plane view with neighbouring structures. The same sections and probability scale as Figure 1 include contours of the lateral and posterior hypothalamus, subthalamic nucleus, zona incerta and red nucleus.

**Supplementary Figure S2.**
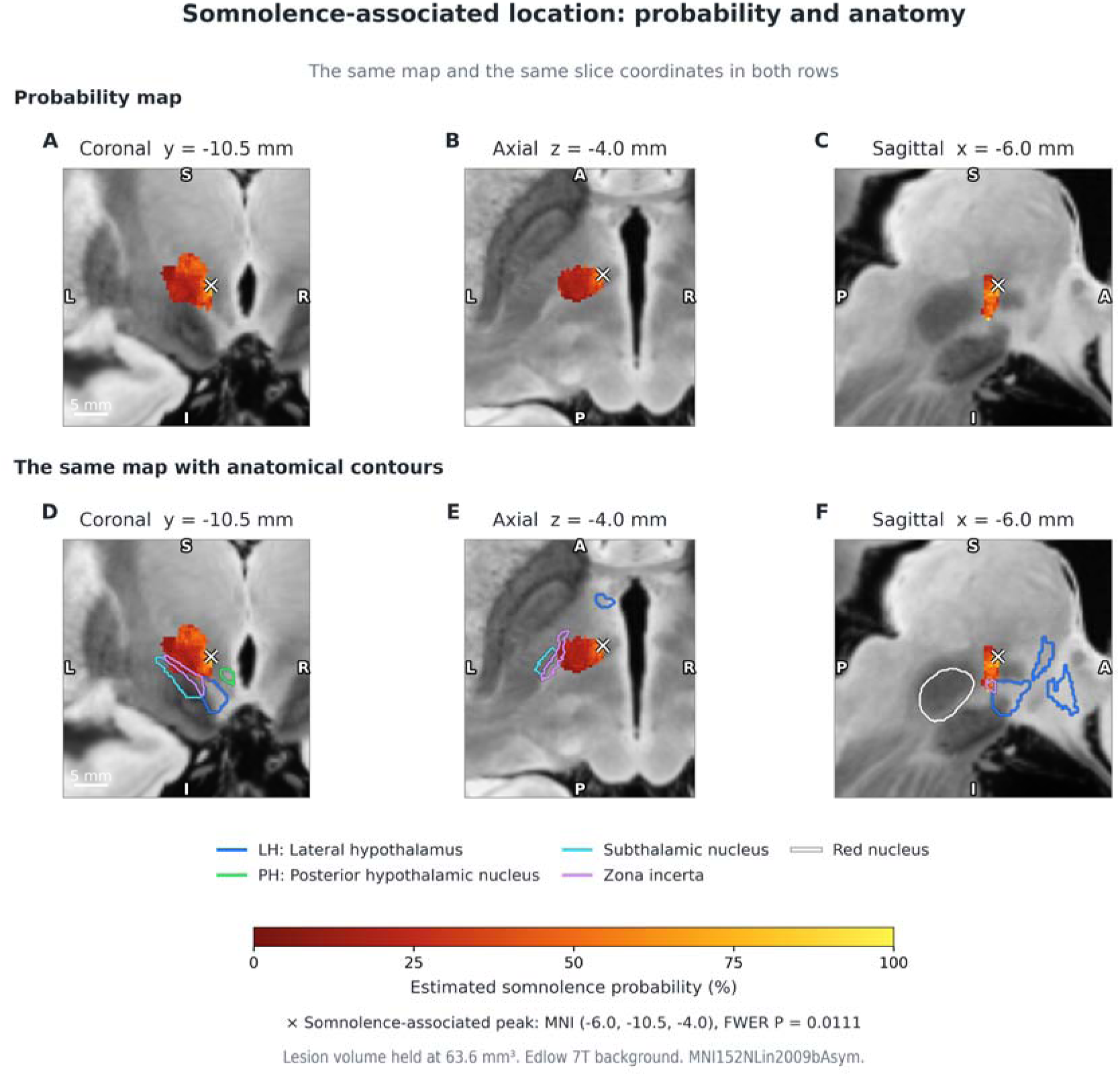
Probability map and anatomical contours. The upper row shows the probability map without anatomical contours. The lower row shows identical planes and values with the five anatomical contours added.

**Supplementary Figure S3.**
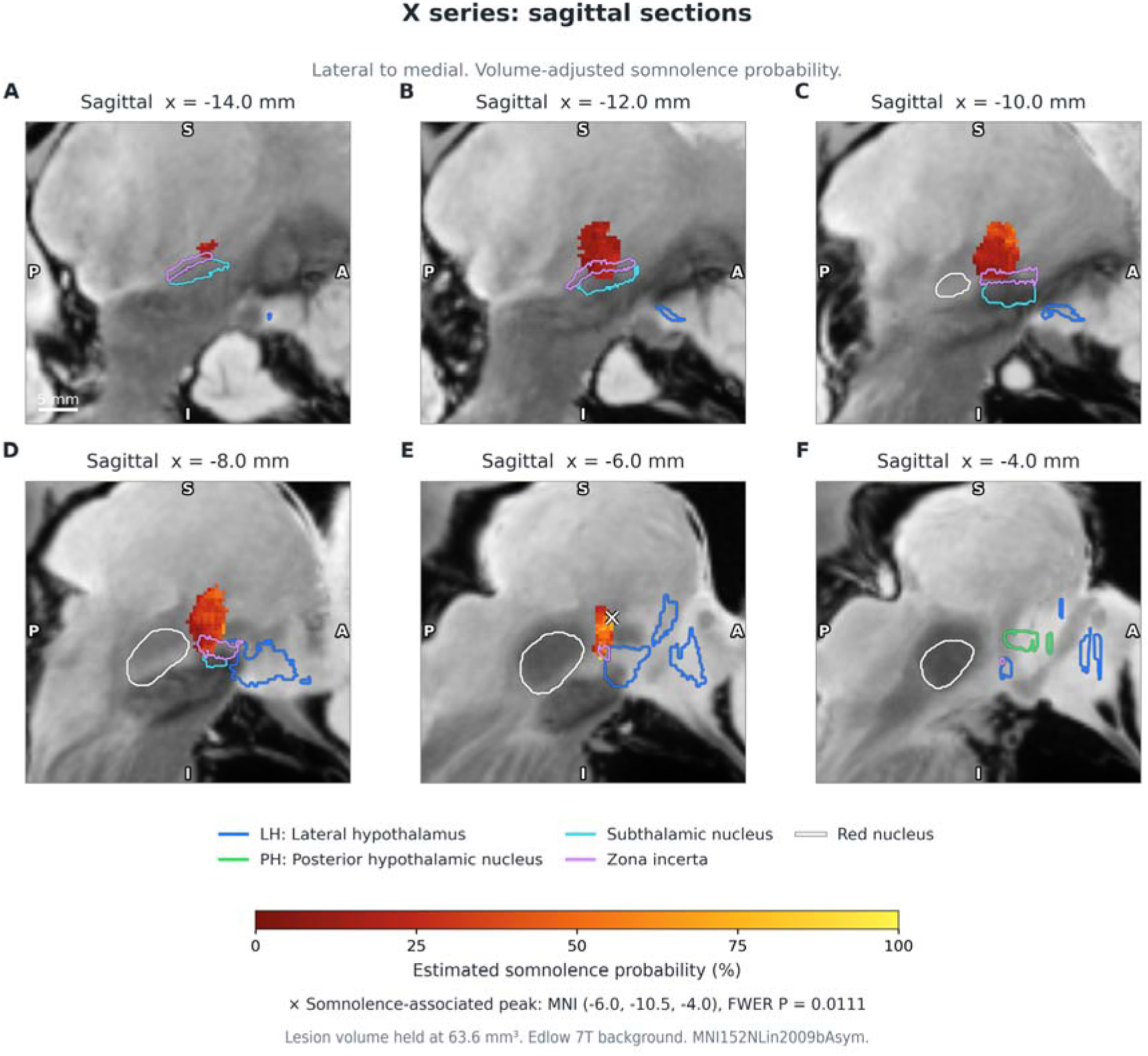
Serial sagittal sections. Sagittal sections run from lateral to medial. The colour scale, anatomical contours and peak marker follow Figures 1 and 2.

**Supplementary Figure S4.**
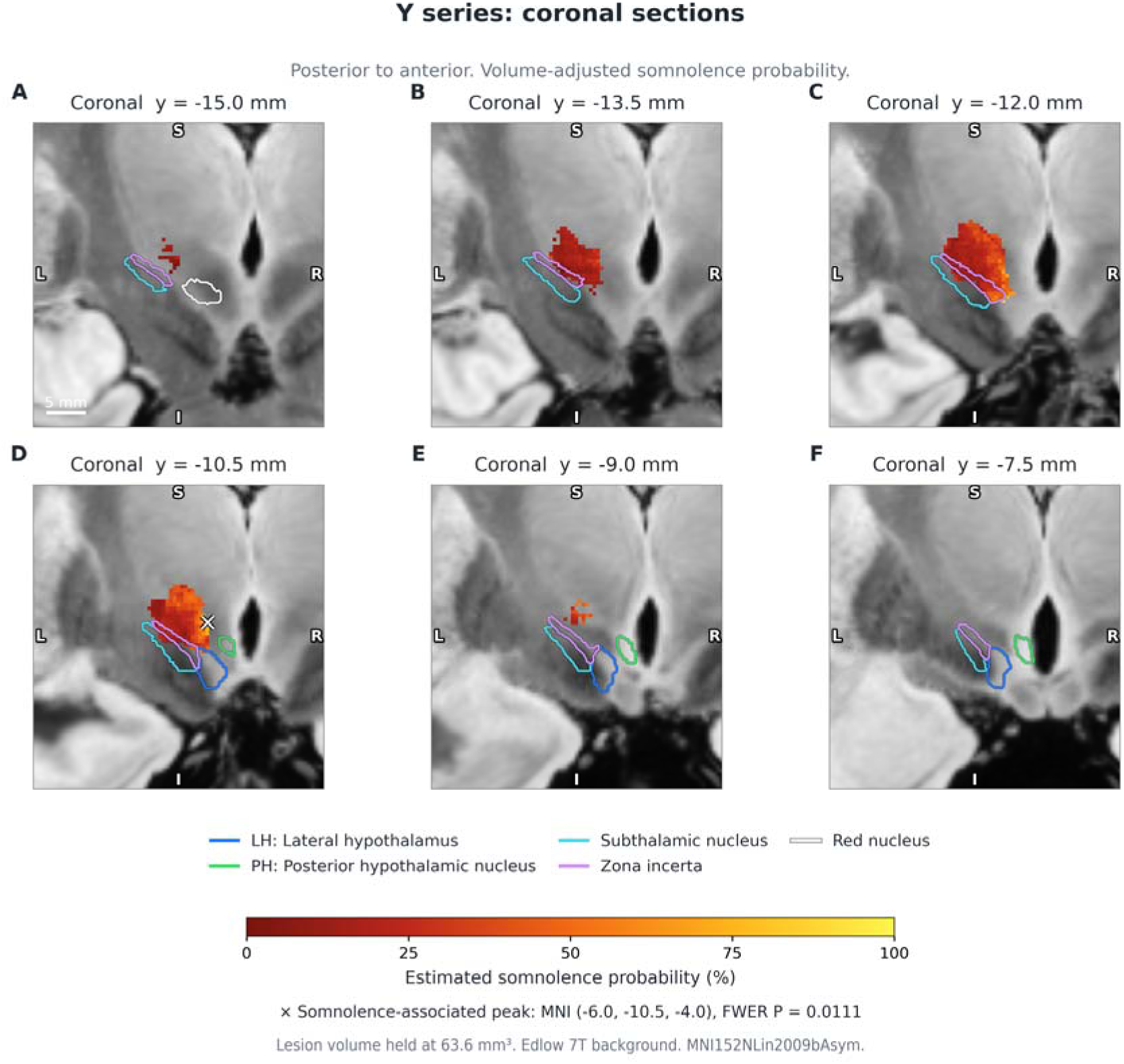
Serial coronal sections. Coronal sections run from posterior to anterior. The colour scale, anatomical contours and peak marker follow Figures 1 and 2.

**Supplementary Figure S5.**
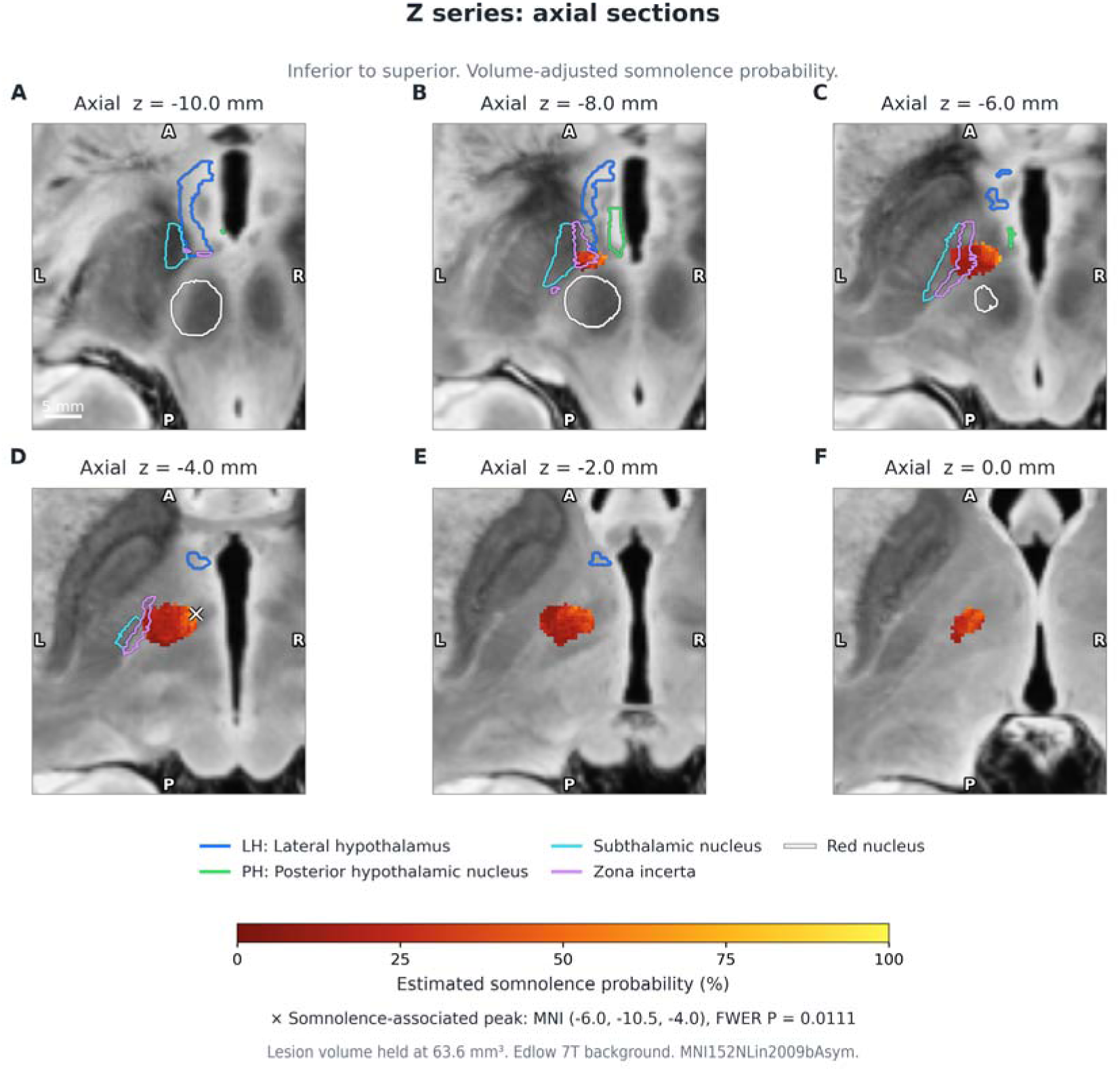
Serial axial sections. Axial sections run from inferior to superior. The colour scale, anatomical contours and peak marker follow Figures 1 and 2.

